# Current Practice and Challenges in Screening for Visual Perception Deficits after Stroke: a Qualitative Study

**DOI:** 10.1101/19013243

**Authors:** Kathleen Vancleef, Michael J Colwell, Olivia Hewitt, Nele Demeyere

**Affiliations:** Department of Experimental Psychology, University of Oxford, Anna Watts Building, Radcliffe Observatory Quarter, Woodstock Road, University of Oxford, OX2 6GG, United Kingdom; Department of Experimental Psychology, University of Oxford; The Oxford Institute of Clinical Psychology Training and Research, University of Oxford

**Author notes:** corresponding author, Twitter: @katvancleef. Twitter: @MichaelJColwell, @NeleDemeyere.

**Keywords:** Visual perception, screening, stroke, assessment, clinical needs, facilitators, barriers

## Abstract

**PURPOSE:** We document current clinical practice and needs in screening for visual perception problems after stroke to inform development of new screening tools.

**MATERIALS AND METHODS:** We interviewed 12 occupational therapists and 13 orthoptists. Interviews were thematically analysed using the Value Proposition Canvas, a model which establishes challenges and facilitators in what people want to achieve.

**RESULTS:** Participants’ understanding of visual perception varied and often included sensory and cognitive deficits. Occupational therapists commonly screened for visual field deficits and hemispatial neglect, while other aspects of visual cognition were rarely assessed. A positive screening result triggered an orthoptic referral. Screening generally occurred during functional assessments and/or with in-house developed tools. Challenges to practice were: lack of time, lack of training, environmental and stroke survivor factors (e.g. aphasia), insufficient continuation of care, and test characteristics (e.g. not evidence-based).

Facilitators were: quick and practical tools, experienced staff or tools with minimal training requirements, a streamlined care pathway.

**CONCLUSIONS:** Screening employs non-standardised assessments and rarely covers higher visual perceptual deficits. We demonstrates the need for an evidence-based visual perception screen, which should ideally be 15 minutes or less, be portable, and require minimal equipment. The screen should be suitable for bedside testing and aphasia-friendly.

## Introduction

Visual perception is the dynamic process of perceiving the environment through sensory inputs and translating the sensory input into meaningful concepts associated with visual knowledge of the environment.(1) Visual perception problems are therefore distinct from sensory visual impairments such as reduced visual acuity, visual field and eye movements.(2) Where sensory visual impairments result from damage to the eye or early visual pathways from the eye to the primary visual cortex, visual perception deficits are attributed to impaired function in later visual processing areas in the occipital, parietal and temporal cortex.(3) Examples of visual perceptual deficits include apperceptive and associative agnosia (object recognition difficulties), prosopagnosia (face recognition difficulties), akinetopsia (difficulties in perceiving motion), achromatopsia (difficulties in perceiving motion), problems in visual memory (remembering what you have seen before), and in visuospatial abilities (e.g. judging distances or spatial relations between objects).(3) Visual inattention or hemispatial neglect is sometimes considered to be part of visual perception,(1),(4) though neuropsychology research attributes this to an attentional deficit.(5) In particular, the presence of preserved perception when attention is stretched to focus on the stimuli, the existence of cross-modal neglect and manipulations of stimulus density on the extent of neglect support the classification of hemispatial neglect as a disorder of attention. (6,7)

Rowe and colleagues reported that 20.5% of stroke survivors with a suspected visual difficulty have visual perception deficits such as visual inattention (14%), visual hallucinations (2.5%), and object agnosia (2.2.%)(8) This study made use of reports by stroke survivors and carers rather than formal assessment (except for visual inattention). With systematic screening with the Rivermead Perceptual Assessment Battery, Edmans and Lincoln identified visual perception problems in 76% of hemiplegic stroke survivors.(9) The discrepancy between prevalence with self-reports compare to neuropsychological assessment suggests that not all visual perceptual problems are picked up based on self-report. This means many stroke survivors are discharged without the appropriate rehabilitation or adjustments in their home environment or care packages.

Under-diagnosis of visual perception problems can severely impact stroke survivors’ quality of life, functional outcome, participation in the community, independence and pose substantial risk. (10–14) For instance, participation in traffic with visual perception problems can be dangerous and even life-threatening. Risks are also heightened indoors when preparing a meal or in perceiving trip hazards. Better diagnosis of visual perception difficulties after stroke will allow better care planning and substantially impact stroke survivors’ life.

The first step toward better diagnosis is an in-depth understanding of the clinical reality and the reasons behind under-diagnosis. We need to understand what the challenges are in screening for visual perceptual difficulties after stroke. This can then inform the development of solutions in the future.

In the current study, we aimed to conduct an in-depth exploration of current clinical practice, challenges and facilitators of screening for visual perception problems after stroke. To achieve a rich understanding of these issues, we performed semi-structured interviews with orthoptists and occupational therapists. These professionals are most commonly involved in visual perception screening after stroke in the United Kingdom’s National Health Service.

## Materials and methods

### Participants

All participants were recruited via opportunistic and snowball sampling. Invitations for the interviews were sent out through (i) email to the British and Irish Orthoptic Society Stroke and Neuro Rehab Clinical Advisory Group, (ii) Twitter, (iii) a sign-up sheet at a conference poster and (iv) informal face-to-face conversations at the United Kingdom Stroke Forum 2018. Participants met inclusion criteria if they were working in the National Health Service as an occupational therapist or an orthoptist, and were involved in the assessment of visual perceptual problems after stroke. The participants were informed about the aims of the study, the organisations running the study (University of Oxford and North Bristol NHS Trust), and the planned outcomes and dissemination. The project was reviewed and approved by the Patient Safety Assurance & Audit Service at NHS North Bristol Trust as a Clinical Effectiveness study (CE45999). All participants gave verbal consent.

### Interviewer’s training

All interviews were conducted over the phone by the first author, KV, who holds an MSc in Clinical Psychology and a PhD in Experimental Psychology. At the time of the research study KV was working as a postdoctoral research fellow at the University of Oxford. KV had nine years’ experience as a researcher, including one year as a qualitative researcher. Her formal training in qualitative research included an undergraduate course in qualitative research methods at the University of Leuven which included a research project. In preparation for the current project, she was trained and supervised by senior author, OH. OH is a clinical psychologist who has worked predominantly as a qualitative researcher and health care professional within the field of intellectual disabilities. OH has numerous publications within this field, teaches qualitative research methods at the University of Oxford, and has supervised a number of doctoral projects using various qualitative methodologies. KV’s training included reading and reflecting on reference works in qualitative research,(15,16) a question and answer session with an experienced moderator, and guidance during coding and analyses via regular meetings with OH.

### Model

The interview process and qualitative analysis was guided by the Value Proposition Canvas. This model was originally designed to guide product development, but can be applied to different contexts.(17) Central to this model is the principle that development of products or services are best informed by the users. A good product or service takes into account the goals of the user, it reduces the pains a user experiences while trying to achieve their goals, and it provides gains in achieving their goals. For instance a good electric car helps a person to get to work (goal to achieve) without having to fear that the battery will run out (reduce pains) while at the same time provides an attractive design to impress colleagues and family (increase gains). We can apply this model in trying to understand current clinical practice in visual perception screening after stroke (goal), the challenges (pains) it brings, and what facilitates the process (gains).

### Data collection

A semi-structured interview guide was developed by KV in collaboration with an occupational therapist and an orthoptist (table 1). The interview guide was reviewed by an experienced moderator and piloted with one orthoptist and three occupational therapists. The interviews consisted of open-ended questions about current practice in visual perception screening after stroke, and the challenges and facilitators of practice. The interviewer conducted interviews at the participant’s workplace or over the phone in a quiet office with no other people present. The participants took the call in a location of their choice. The location and presence of others at the participant’s side was not recorded. After obtaining participant consent, the interview was audio recorded. All participants were interviewed once. Recruitment was ongoing during data collection and interviews were conducted until data saturation was reached.

**Table 1.**
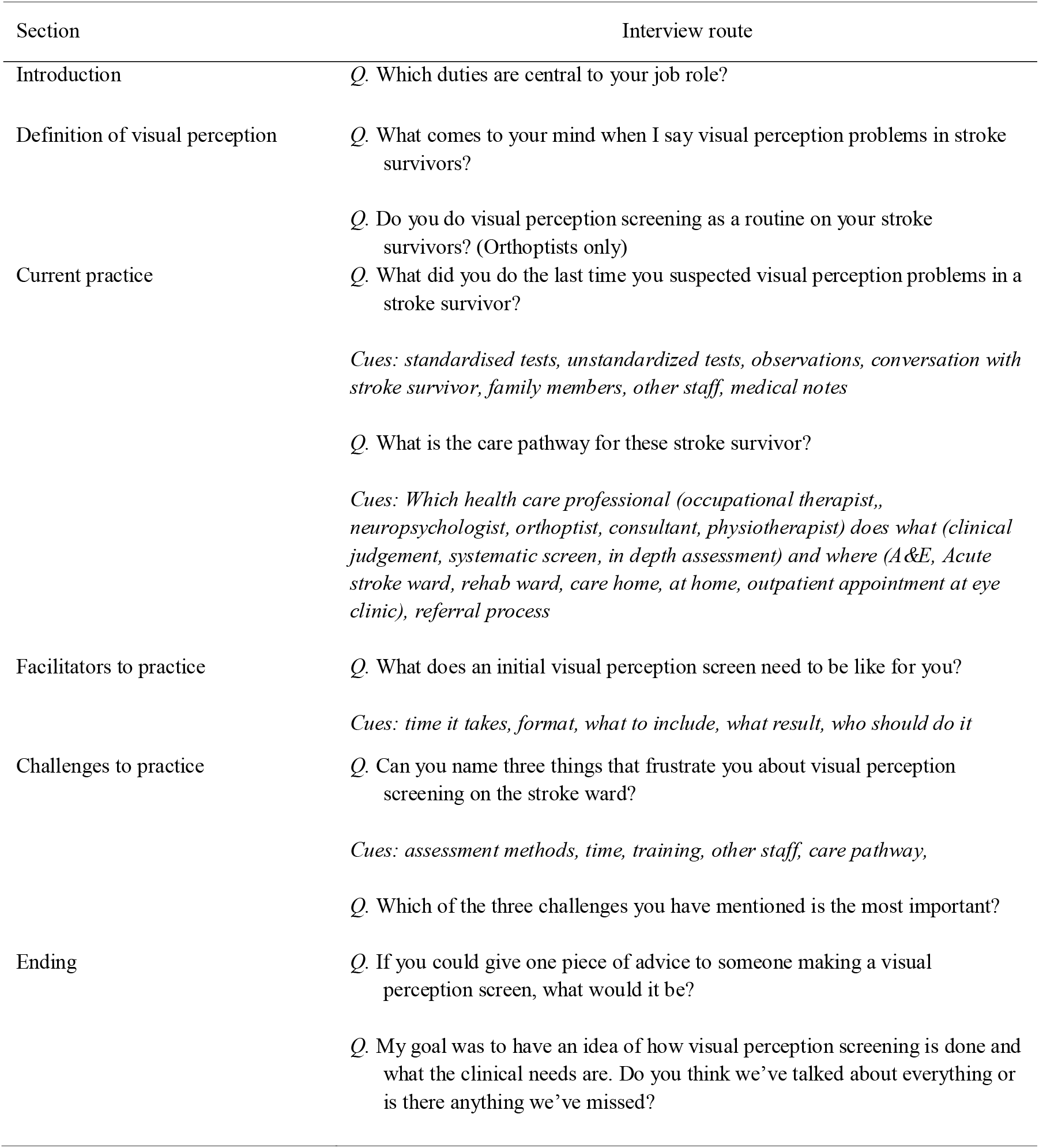
Interview structure

### Qualitative Analyses

As a method of data analysis we used thematic analysis.(18) Interviews were transcribed verbatim by a professional typist. The transcripts were not returned to the participants for feedback. Two researchers (KV and MC) were involved in data analysis. KV was familiar with the data from conducting the interviews (Phase 1 in thematic analysis). The first 15 interviews were checked and coded by KV in NVivo Software (Version 12.0). She developed an initial coding tree (Phase 2 in thematic analysis). Overall themes were set in advance in alignment with the Value Proposition Model: Jobs, Challenges, and Facilitators. The coding tree was reviewed and substantially adapted after coding of five interviews. MC familiarised himself with the data by checking transcripts and reading through interviews coded by KV. Two of these interview transcripts were checked, coded and discussed by both KV and MC to align their coding. MC checked and coded the remaining 10 interview transcripts. The frequency of text extracts coded under each node was visualised with a hierarchical chart of nodes in NVivo to explore overall patterns in the data describing current practice. The use of frequencies in qualitative analysis can be a valuable strategy when used as a complement to an overall process orientation to the research. They can provide evidence for generalizability of the findings between participants, highlight diversity, reveal larger patterns beyond a participant’s immediate experience, minimise cherry picking of the data.(19) This chart shows the frequency of references for each node when participants were talking about current practice in routine visual and visual perception screening. If a participant referred to a node multiple times during one interview, each reference was counted. Only nodes with at least ten references were included in the hierarchical chart of nodes. Overall themes and subthemes were derived from all data by KV (Phase 3 of thematic analysis). This was done in two stages: (1) we performed search queries of coded text for challenges and facilitators of visual perception screening in current practice and (2) read through the resulting text. The themes were subsequently reviewed by reading all answers to the questions under the headings Challenges and Facilitators (see table 1). The information was then synthesised by KV (Phase 4 of thematic analysis). Lastly, themes and subthemes were reviewed, refined and defined by KV (Phase 5 of thematic analysis) and a report was prepared (Phase 6 of thematic analysis). This report was sent out to the participants for validation. Ten out of 25 participants replied and nine agreed with the thematic analysis. One participant pointed out that participants’ understanding of visual perception was underexplored. We added a results section on this topic posthoc.

## Results

12 occupational therapists and 13 orthoptists from 16 health care organisations in England took part in the study. Characteristics such as gender, years of experience and main clinical setting of each participant are reported in table 2. All participants worked in England, and were based across the East Midlands (n=1), East of England (n=1), Greater London (n=2), North East (n=3), North West (n=4), South East (n=6), South West (n=4), and Yorkshire and the Humber (n=4). The interviewer had no personal relationship with any of the participants. Two orthoptists received referrals from the same acute stroke unit where the first author recruits stroke survivors as participants for other studies (pseudonyms cannot be disclosed to ensure anonymity). One of the participants was also involved in the development of the interview guide. All but one participant (Jessica) agreed to be audio recorded. For Jessica, notes were made during and immediately after the interview. Interviews lasted between 16 and 46 minutes with an average of 27 minutes. For Christopher, the interview was split over two consecutive days.

**Table 2.**
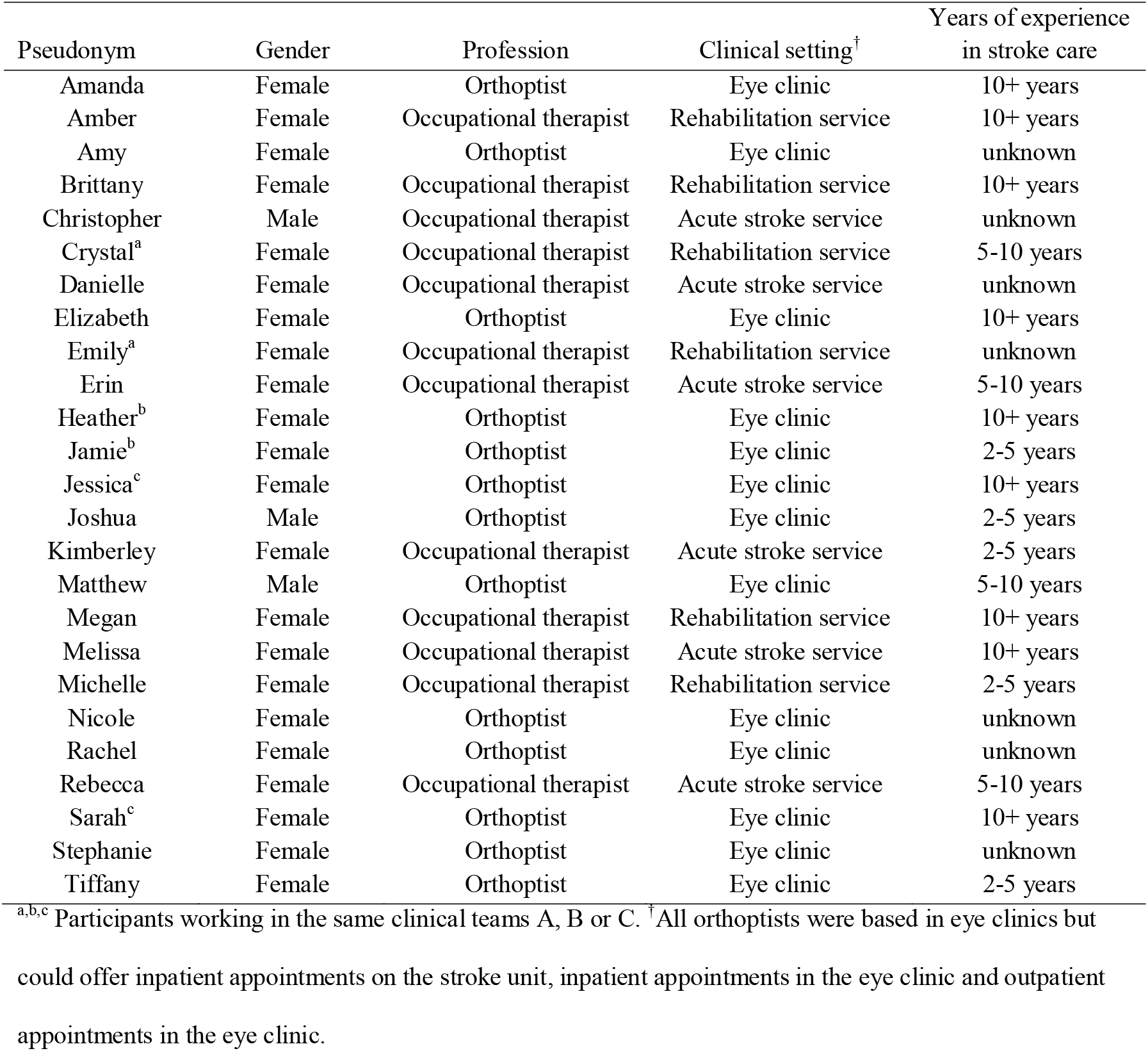
Participant characteristics

### Lack of clarity around visual perception

The interviews highlighted that participants’ understanding of visual perception differed from textbook definitions of visual perception. Visual perception is defined as a dynamic process of translating sensory visual information into meaning percepts(1). These percepts can subsequently be linked to higher cognitive functions like visual memory for recognition (ventral or what stream) and praxis for visually guided actions (dorsal or where stream)(3). Visual perception must be differentiated from low level sensory visual functions like acuity, visual fields, and ocular movements, and from higher cognitive processes like (visual) memory or (visual) inattention, although some authors consider visual inattention to be part of visual perception. Our participants seemed to have a broader understanding of visual perception and often did not differentiate between sensory and higher cognitive functions. Participants typically mentioned the following examples when asked about what they consider visual perceptual problems (in descending order of frequency): visual inattention, agnosia’s for objects, faces, letters, or shapes, hallucinations or Charles Bonnet syndrome, difficulties in depth perception, visual field deficits like hemianopia, and spatial vision (e.g. navigating and perceiving space around them).

Participants’ understanding seemed to vary widely but we observed general trends in the different professions. Orthoptists seemed to differentiate visual perception difficulties from low level sensory visual impairments, but considered visual inattention the prime (and only) example of visual perception difficulties after stroke.

> “[*Interviewer: What comes to your mind, when I say visual perception problems in stroke patients?*] So I would probably say visual neglect, ignoring one side of their world when the other side is presented to them as well, is the main definition of it. That it’s decreased awareness of that side of their vision and their external world as well as themselves as well, on that side. *[Interviewer: Anything else, that falls under that term for you?]* No, not that I can think of, off the top of my head, no.” (Tiffany, Orthoptist in an eye clinic)

In addition to visual inattention, occupational therapists mentioned impairments that are typically (but not exclusively) related to low level sensory vision like contrast sensitivity, hemianopia, and depth perception. They seemed to interpret visual perception more broadly.

> “A range really. So we would look at visual perception as things like depth perception, … recognition of objects, … and any kind of visual inattention or neglect, that sort of thing… But yes, that’s it basically.” (Danielle, Occupation Therapist in an acute stroke service)

These observations are relevant in interpreting the subsequent results. Although the interview questions asked for experiences with visual perception assessment after stroke, the results below should be interpreted as potentially referring to all or any of the following: sensory visual impairments (for occupational therapist participants), visual perception impairments (for all participants), and visual inattention (for all participants).

### Current Practice

A hierarchical chart of nodes is presented in figure 1. The figure illustrates that participants most often mentioned occupational therapists working in acute stroke or rehabilitation services when discussing screening for visual neglect, visual field deficits, and ocular movements. For screening they referred to in-house developed screening tools, standardised tests or observations in functional tasks like dressing and making a cup of tea.

**Figure 1.**
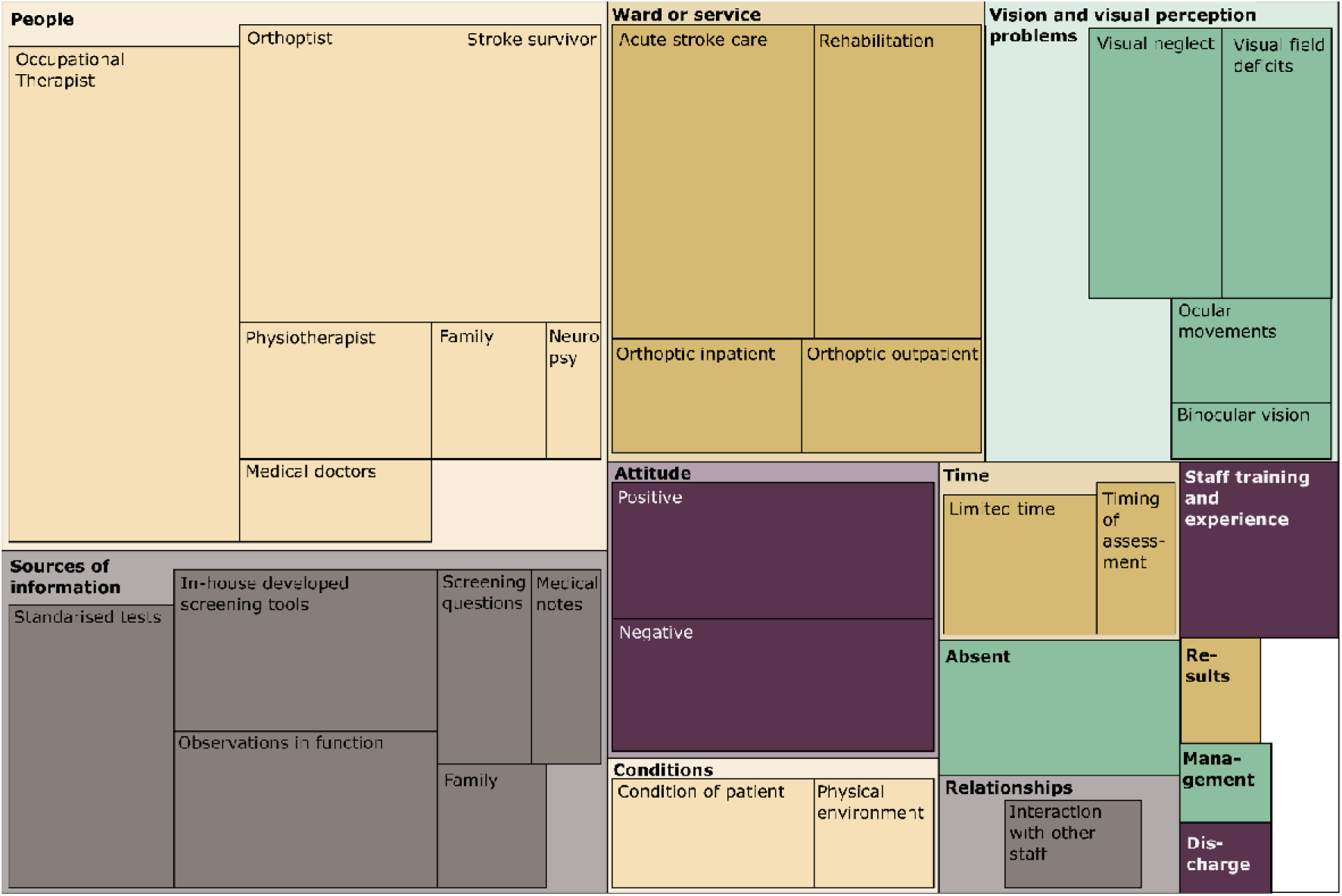
Hierarchical chart of nodes showing the frequency of references for each node. The size of a box reflects the frequency of references for each node with larger boxes demonstrating more frequency mentioned nodes. Parent nodes are shown in lightly shaded boxes, child nodes are shown in darker shaded boxes within their parent’s box. The colours are added for easy distinction between different parent boxes but have no further meaning. Text is filtered for extracts where participants were talking about current practice in routine visual and visual perceptions screening. Only nodes with at least ten references are shown.

> “In terms of the visual field, we do the standard, you know, wiggle your fingers test to see how people respond. We do the tracking tests. Otherwise it’s mainly through observing people in functional tasks what they’re doing Through that you can often realise if they’re having problems with their visual perception because they’re missing things or not seeing things.” (Crystal, occupational therapist in a stroke rehabilitation service)
>
> “I’d rather do it in function. It’s more meaningful especially with stroke patients because it makes more sense. Rather than getting them to balance cubes on each other or, you know, I can say ‘Can you find the toothpaste?’ “(Megan, occupational therapist in an acute stroke service)

Most participants mentioned that the service they work within has a referral pathway in place: if problems are suspected following screening by the occupational therapist, a referral is made to orthoptics. A stroke orthoptist will then see the stroke survivor as an inpatient on the unit or as an outpatient after discharge from the hospital.

> “It’s really as and how other health professionals have identified them, then an orthoptist will go up to try and assert a diagnosis and then give them the rehabilitation options.” (Amy, orthoptist in an eye clinic)

### Challenges

Themes around challenges in visual perception screening are presented in table 3. We identified five major themes and 13 subthemes.

**Table 3.**
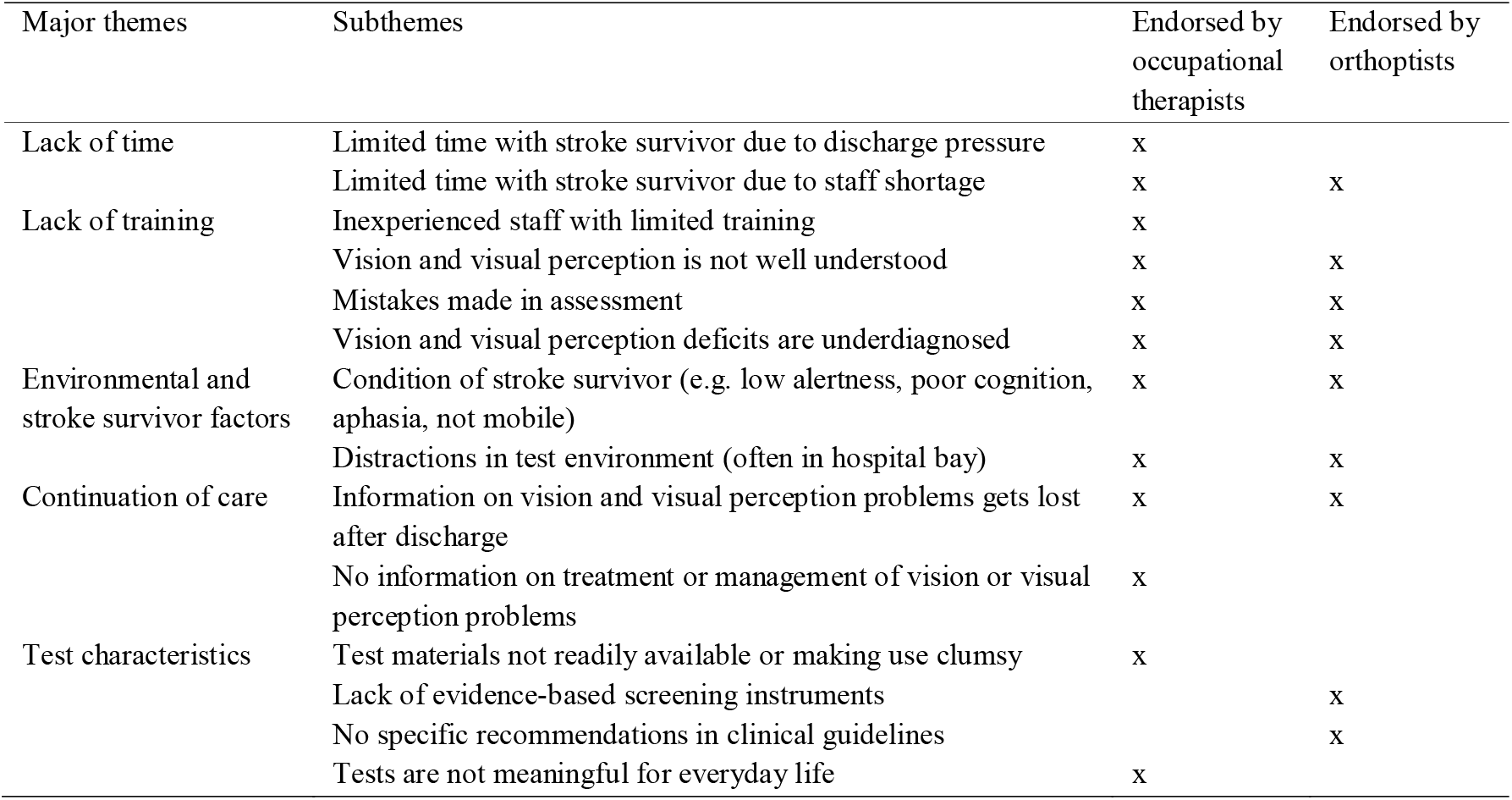
Challenges in visual perception screening

#### Lack of Time

Occupational therapists and orthoptists both reported that time pressure makes visual perception screening difficult. They mentioned that because of understaffing they spend little time with stroke survivors or experience delays in seeing them following referral. In addition, occupational therapists report that stroke survivors are discharged from an acute stroke unit as soon as medically safe and a suitable discharge location has been organised. For survivors with milder strokes, discharge is reported to happen within 1-2 days, giving very little time for therapists to complete their assessments.

> “Well one *[challenge]* is just logistically and organisationally, just in terms of caseload and time to hand. We are having to prioritise more direct functional assessments in terms of establishing readiness for discharge.” (Christopher, occupational therapist in an acute stroke service)
>
> “There’s not a lot of OTs who are actually for *[delivering]* the therapy, so time-wise we haven’t got a lot of capacity at times.” (Danielle, occupational therapist in an acute stroke service)
>
> “Lack of time is always going to be frustrating knowing that you’ve got a clinic here and you can’t get up to the ward and there’s an urgent stroke survivor.” (Heather, orthoptist in an eye clinic)
>
> “Erm… it sounds awful doesn’t it when you’re constantly having to slim down what you do.” (Melissa, occupational therapist in an acute stroke service)

#### Lack of Training

Training is another theme that frequently emerged across both professions. Visual perception difficulties following stroke appear to be only briefly covered in the undergraduate education of orthoptists and occupational therapists, and is mostly learned about post-qualification through on-the-job-training. Many occupational therapists mentioned that they rotate between services, leaving little time for them to be trained in many aspects of stroke care. In particular, they said that this lack of experience made them feel uncertain when it comes to vision and visual perception deficits following stroke. Inexperience and limited training seemed to be an important hurdle for occupational therapists in visual perception screening. Because vision and visual perception was not well understood by the staff (see section ‘Lack of clarity around visual perception’), participants reported that mistakes in assessments are commonly made, leading to a potential under-diagnosis of visual perception deficits.

> *“[It]* can be a bit frustrating … lots of the occupational therapists are on a rotational job. So getting them *[occupational therapists*] trained up *[is difficult]*, I think they *[occupational therapists]* do find vision scary Vision, it’s the scariest thing about stroke.” (Stephanie, orthoptists in an eye clinic, using ‘vision’ as a general term to cover a wide range of sensory and perceptual visual functions)
>
> *“[Visual perception is]* one of those things that is perhaps not widely understood. think unless you’ve got a unit with a special interest in stroke and you’ve taken the time to look into that. then it’s perhaps under-diagnosed as well, so yeah I think it’s recognition that it’s a potential problem.” (Amanda, orthoptist in an eye clinic)
>
> “The key is really, somebody in the hospital to be able to deliver the training and the therapy staff to actually take on board the training and to actually deliver it.” (Sarah, orthoptist in an eye clinic)
>
> “I think it probably is underreported; the amount of incidents of perceptual problems post stroke.” (Rachel, orthoptist in an eye clinic)

#### Environmental and Stroke Survivor Factors

Participants reported that visual perception screening is limited by the environment in which they take place. They said that in acute stroke care, most assessments are done in a hospital bay with several beds. Even if curtains can be drawn around the stroke survivor’s bed, distractions were reported to be numerous: sounds from neighbouring beds, interruptions for timed caring needs, distractions from medical equipment around the bed and presence of personal objects on a small table in front of the stroke survivor (spectacles, drinks, food, tissues, personal care items, newspaper, get well cards, crossword book, pen, etc.) can be visually distracting and interfere with visual perceptual assessment.

> “Frustrating for us is … that we .. depending on which unit we are *[on]* or where we are, it can be quite difficult to do *[our assessment at]* the bedside if there is a lot *[of]* distractions going on; if there is no quiet space to take the patient.
>
> Obviously, yes, we can draw the curtains but it’s sometimes more the noise levels than everything. They get easily distracted. That makes it quite hard actually to do an assessment.” (Amber, occupational therapist in a stroke rehabilitation service)
>
> “It’s *[the]* environment. Often *[it]* doesn’t lend itself to a nice quiet space where patients can attend.” (Danielle, occupational therapist in an acute stroke service)

Participants also mentioned the condition of the stroke survivors as a limiting factor to visual perception screening. Both professions reported that tests and tools are often not suitable for stroke survivors with poor alertness, poor cognition, aphasia or severe weakness in their upper limbs. For instance, stroke survivors may not able to answer a question on whether their vision has changed because they developed aphasia following stroke. Similarly, copying and cancellation tasks to assess visual neglect were reported to be unsuitable for a stroke survivor who can no longer hold a pen.

> “Another component would be around the client or the patient themselves in terms of factors that might work against screening: so attention, concentration, fatigue, the ability to deliver a verbal response in terms of aphasia or dysarthria difficulties. That has significant impact on our ability to undertake formal testing of whatever nature.” (Christopher, occupational therapist in an acute stroke service)
>
> “Our patients who either are fatigued quite quickly, or are medically not well enough to do a lot of taking down to the therapy department, or their attention span is quite limited, or they’ve got a weakness in their upper limb that makes it more difficult to complete the tests.” (Danielle, occupational therapist in an acute stroke service)

#### Continuation of Care

A further concern of the participants was the follow-up process after screening for visual perception deficits. They highlighted that quite often the information from screening is not passed on to the next care team. Therefore, even if visual perception problems are picked up in an acute setting, no follow-up assessments are carried out or no rehabilitation is provided in their experience. With respect to rehabilitation, occupational therapists found it frustrating that few treatment options are available. They reported that little information and few guidelines are available on how visual perception problems can be treated or managed. Some noted that the lack of options for rehabilitation reduced their motivation to screen for visual perception problems.

> “Yes, and it [*the discharge summary*] is very medical because the occupational therapists or myself don’t like the discharge summaries We do all this information and then it gets lost ….Yeah, it’s not part of our discharge summary in enough detail, so when they go somewhere new which could be two or three weeks later … they don’t have that information.” (Matthew, orthoptist in an eye clinic)
>
> “It might be because it [*visual perception deficits*] is limited in treatment … that’s why it’s not the focus. So … it would be more useful to spend time during my assessment speaking to them about adapting to their field loss and giving them a prism or occlusion for their double vision.” (Rachel, orthoptist in an eye clinic)

#### Test Characteristics

Both professions highlighted limitations in the characteristics of the available tests for visual perception. Occupational therapists more frequently reported to be hindered by practical limitations and the unknown impact on daily life. With respect to the practical limitations, they referred to a large number of test materials like stimulus booklets, stopwatch, cubes, etc. needed for assessment or too many loose pages. With respect to the unknown impact on daily life, they referred to stimuli and tests that were thought to be limited in ecological validity. They considered the tests too abstract with no clear link to the implications for everyday tasks.

> “We have our little screening tool. It’s a few pages, just to prompt us That’s all right but then … everything else you are searching together from different kits. … That’s a little bit frustrating.” (Amber, occupational therapist in a stroke rehabilitation service)
>
> “There’s an awful lot of bits of paper and a bit of faffing about and preparing to get it all ready before we actually go out to see the patients.” (Melissa, occupational therapist in an acute stroke service)
>
> “We use Rivermead Perceptual Battery. But to be fair, I don’t tend to use it as often because I’d rather do it in function It’s more meaningful, especially with stroke patients because it makes more sense. Rather than getting them to balance cubes on each other or, you know I can say “Can you find the toothpaste? Can you take the lid off? Can you brush your own teeth?” and then it just makes more sense. And then it’s easier for them to identify *[a]* goal.” (Megan, occupational therapist in a stroke rehabilitation service)

Orthoptists, on the other hand, seemed to emphasize more the lack of evidence and consequently the lack of specific clinical guidelines on how to assess visual perception.

> “If you see the national guidelines for visual problems after stroke, they have a really small section on vision with orthoptics but we don’t really have anything in it that’s specific to what we should be screening them with and what we should be treating them with. So even though we’re using evidence-based practice, the evidence sometimes isn’t great and I think that’s probably why it’s not in the guidelines because there’s not really good evidence to support it. But sometimes you’re seeing a patient and you’re not entirely sure if you’re doing the right thing because you’re maybe giving them information based on clinical experience, what other people have been doing but you’re not really sure if that is the best thing to be doing or could you be doing something else with them.” (Amy, orthoptist in an eye clinic)

### Facilitators

The facilitators to practice that participants reported, largely mirrored the challenges reported above (table 4). We identified three major themes and 12 subthemes.

**Table 4.**
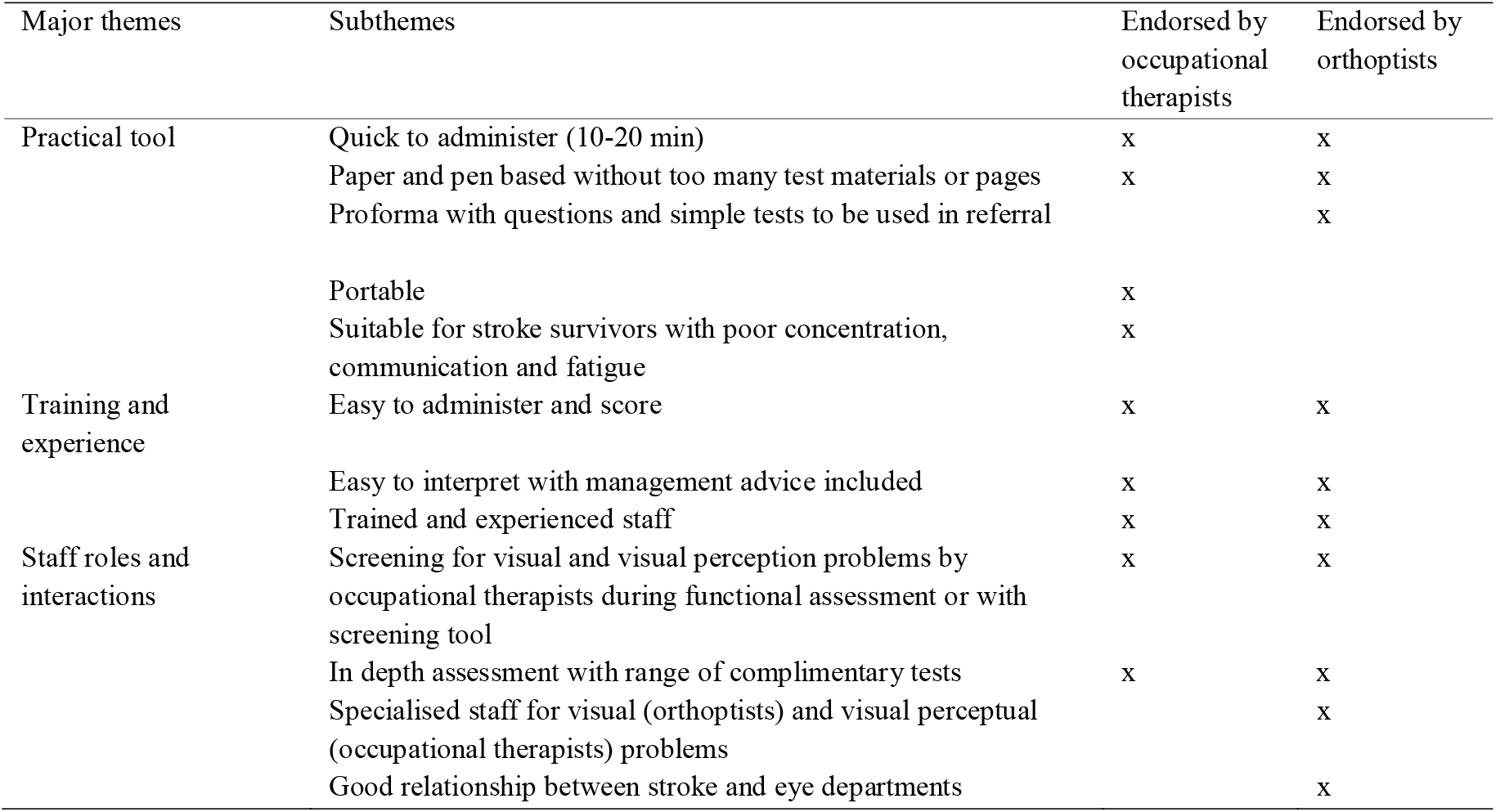
Facilitators to visual perception screening

#### Practical Tool

On a practical level, participants preferred a tool that can be administered in approximately 10-20 minutes, so it can fit in with busy every-day practice and current staff levels. They also favoured a tool that consists of few test materials and pages to prevent test materials getting lost or mixed up during testing. According to participating orthoptists, the tool should have a 1 to 2 page proforma. They reported that it should include questions to ask the stroke survivor and simple quick tests for a range of functions. They suggested that the proforma could subsequently be used in a referral for orthoptic input. To allow bedside testing or testing in the community, participants preferred portable tools. Lastly, many participants highlighted that a tool should ideally be suitable for all stroke survivors, such as those with aphasia, poor concentration, poor cognition or severe fatigue.

> “It needs to be something that’s fairly quick and easy to administer.” (Danielle, occupational therapist in an acute stroke service)
>
> “Just pen and paper, quite simple or a few things to hand that will fit in something that’s quite portable and mobile, I think [*that*] would be handy, yes.” (Brittany, occupational therapist in a stroke rehabilitation service)
>
> “Reasonably short … for logistical [*reasons*]: … our time and patient concentration time, fatigue in the more acute stages. I think really half an hour is the maximum time.” (Christopher, occupational therapist in an acute stroke service)
>
> “It would have to be a list of standardised questions: Are they able to look between two points specifically? … I guess the visual perception question would be if they’re following a moving object? … You know are they able to identify their friends or their family? Can they recognise themselves in a photo?” (Rachel, orthoptist in an eye clinic)

#### Training and Experience

Orthoptists and occupational therapists agreed on the importance of training and experience for robust visual perception screening. The presence of experienced colleagues or training by other staff members was seen as very advantageous. Given the limited training in visual perception highlighted above, a tool that is easy to administer and score was preferred by our participants. Similarly, they reported that results should be easy to interpret, and should provide clear guidance for management of the deficit (if present).

> “If it was something that was quite self-explanatory then anybody could do it and therefore anybody can interpret it, then that would be much clearer.” (Amy, orthoptist in an eye clinic)
>
> “Just easy to administer and easy to score. … So making instructions clear and easy to understand would be good.” (Emily, occupational therapist in a stroke rehabilitation service)

#### Staff Roles and Interactions

Occupational therapists reported that their preferred practice is to screen stroke survivors for vision and visual perception problems during their functional assessments (e.g. washing, making a cup of tea) or via a screening tool. Both professions mention that if a problem is flagged, more in-depth assessment can then take place.

> “Whereas since we’ve set up a proper vision screening it just means that things have been a lot more efficient and that’s made a much better service because they’re not having to see a lot of people that don’t need to be seen.” (Nicole, orthoptists in an eye clinic)
>
> “So, I’m happy with the main basic screen to begin with, and if it digs something up then we’ll dig it in some more detail. I think that works quite well.” (Matthew, orthoptists in an eye clinic)

If the problem concerns visual perception difficulties, an occupational therapist is seen by our participants as the most appropriate professional to follow it up. If the problem concerns sensory vision, a referral to an orthoptist was the preferred practice in our sample. Good relationships between stroke occupational therapists and orthoptists in eye clinics were frequently mentioned as invaluable.

> “I think the key is having the staff, the occupational therapist and the physio staff actually take that on board.” (Sarah, orthoptist in an eye clinic)

## Discussion

The aim of this qualitative research was to gain a deeper understanding of current practice in visual perception screening after stroke, the challenges faced by health care professionals. In our participants’ stroke care services, occupational therapists most often screened for visual and visual perceptual difficulties. If visual difficulties like hemianopia, double vision or reduced acuity, are found, a referral to an orthoptist is made for an in depth assessment. This referral strategy corresponds to an earlier report of 76.7% of stroke units having access to an orthoptic stroke service.(20) If visual neglect is suspected, occupational therapists will often follow this up themselves with a range of tasks like cancellations, copying, line bisection, etc. The biggest challenges faced by our participants were lack of time and training. Facilitating factors mentioned were simple, easy to use instruments and good relationships between stroke and eye departments.

The qualitative approach of our study allowed us to gather views directly and in depth. Instead of reporting frequency of views (e.g. how many health care professionals report staff shortage), we focus on how central certain views are to a person’s experience. Our sample covers the two main professions involved in visual perception screening after stroke.

National Clinical Guidelines recommend that every stroke survivor who appears to have perceptual difficulties should have an assessment with standardised measures.(2) As is evident from our interviews, only a small proportion of stroke survivors receive an assessment with standardised measures for visual perceptions difficulties, and screening is not routinely done. The means of assessment of visual perception deficits varied widely between participants. Orthoptists will often use standardised tools in their assessments, though these are not necessarily validated with stroke survivors.(21,22) Overall, standardised assessments were rarely mentioned by our participants. Occupational therapists expressed preferring assessment during functional tasks like washing and dressing and assessment with in-house developed screening checklists. This may be due to a lack of a validated screening tool for a wide range of visual and visual perceptual deficits that can be used by any health care professional.(21) The diversity in tools that we observed in our group of English participants contrasts with the reported use of standardised tools in an Australian population where the Occupational Therapy Adult Perceptual Screening Test and the Loewenstein Occupational Therapy Cognitive Assessment are most commonly used to evaluate visual perceptual difficulties.(22)

McCluskey and colleagues investigated the barriers and enablers for following recommended practice in stroke care.(23) In line with our findings the occupational therapists they interviewed reported the condition of the stroke survivors as a barrier for standardised assessments. Many tests are not designed and not suitable for stroke survivors with aphasia and/or dysarthria. Occupational therapists in their study also mentioned the lack of training and knowledge, specifically for visual neglect rehabilitation. Time pressure and fluctuating staff levels meant occupational therapists in their study prioritised assessments and interventions which would produce the best clinical outcome. A lack of time to see stroke survivors was also mentioned by their orthoptists. However, in our study this referred to the lack of time to see all referred stroke survivors before they were discharged, while in the study by McCluskey, the orthoptists referred to the lack of time to treat stroke survivors. The standardised assessments used by Australian occupational therapists, Occupational Therapy Adult Perceptual Screening Test and the Loewenstein Occupational Therapy Cognitive Assessment, require training and take 20-45 minutes to complete. Given the frequently reported lack of training and time, it may appear that the standardised tools recommended by clinical guidelines are not adapted to the clinical reality of the National Health Service in England. Although McCluskey’s study was limited to one Australian hospital and only involved 5 occupational therapists and 2 orthoptists, all working in the same service and circumstances, the barriers are remarkably similar to what our 12 occupation therapists and 13 orthoptists reported experiencing in 16 English healthcare services. The stroke survivor’s condition, lack of time and staff seem to be universal barriers for providing evidence-based stroke care across domains, not just visual perception screening. Also, the lack of information on treatment or management of vision or visual perception problems that occupational therapists mention as a barrier is not surprising given the very limited evidence for treatment and management options.(24)

The first challenge exposed by our study is the variation between participants in their understanding of constitute visual perception problems after stroke. This led to participants answering interview questions with different reference frameworks. When sharing the challenges of visual perception screening in their clinical practice, some might have been considering challenges in just screening for visual inattention difficulties while others considered a broad range of sensory and cognitive visual impairments. To maintain an equal relationship between participant and interviewer, and an open non-judgemental environment to freely share their experiences, participants’ definition of visual perception was not challenged during the interview. A disadvantage of this approach is that it is unclear whether certain themes apply specifically to one aspect of visual and visual perception screening or to the whole range of vision related difficulties after stroke.

The second challenge lies in our sample size. Our sample, though the largest for a qualitative study in this topic, is not representative for the whole population of health care professionals involved in visual perception screening after stroke. We limited ourselves to occupational therapists and orthoptists as they are most commonly involved in visual perception screening in England. Experiences might be very different in clinical settings without an orthoptic department or with more involvement of neuropsychologists in stroke care. In addition, voluntary participation and asking our participants to commit 30 minutes of their time might have induced a bias for allied health professionals with a keen interest in the topic. We have possibly missed opinions from people with a more negative or neutral attitude towards visual perception screening. In addition, newly qualified health care professionals with limited experience in stroke care might not have felt confident enough to share their opinions on the topic. Lack of confidence on this topic with junior staff was brought up by several of our participants. However, the aim of this qualitative research was not to generalise findings to the population, but rather to provide an in-depth exploration of the topic to generate hypotheses. We are following this research up with a large scale survey for all health care professionals involved in visual perception screening in the United Kingdom and Republic of Ireland. In the recruitment to this survey study, we are emphasizing that we welcome the opinions of heath care professionals with all levels of experience.

A third potential limitation is that the interviewer was the principal investigator of the study and was therefore not independent. Unconscious bias of the interviewer might have guided the participants’ answers. We have tried to minimize this by using a detailed interview guide that included a list of cues that could be given. In addition, all interviews were recorded and a second coder was involved.

Our study limits itself to health care professionals’ perceptive on visual perception screening after stroke. We have not explored experiences by stroke survivors, although they can be substantially different. Also, we have not investigated the management of visual perceptual problems after stroke of their impact in daily life as is previously reported for sensory visual problems. (25–27)

The current research highlighted that a clear and consistent definition of visual perception should be provided to orthoptists and occupational therapists. More training is needed on the assessment (and management) of visual perceptual screening. This is preferably provided via professional organisations to ensure consistency and in line with national guidelines(2,28). This will raise awareness and reduce insecurity experienced by junior staff members. Establishing good relationships between staff at stroke and eye departments is strongly recommended. We suggest this could be achieved in the form of knowledge exchanges or via a re-evaluation of the existing referral system. Lastly, a quick and portable visual perception screening tool that is easy to administer, score and interpret would highly benefit both staff and stroke survivors if it is inclusive for stroke survivors with aphasia, motor impairments and cognitive problems. The tool should be evidence-based and self-explanatory to use. Such a tool is currently not available. Our research provides test developers with the requirements of occupational therapists and orthoptists for such a screening tool to be clinically useful and thereby increase the likelihood of adoption.

## Data Availability

Given the small group of stroke orthoptists and stroke occupational therapists in England, sharing the interview transcripts would potentially disclose participants' identity.

## Acknowledgments

We would like to express our sincere gratitude to all the participating allied health professionals who gifted their time. In addition we would like to thank Philip Clatworthy and Claire Murray for facilitating this project. This work was supported by the Stroke Association under TSA PDF 2017/03; and under TSA LECT 2015/02.

## References

1. Bouska MJ, Kauffman NA, Marcus SE. Disorders of the visual perceptual system. In: Umphred DA, editor. Neurological Rehabilitation. 2nd editio. St. Louis, MO: The C. V. Mosby Company; 1990. p. 705–40.

2. Intercollegiate Stroke Working Party. National clinical guideline for stroke. 2016.

3. Kolb B, Whishaw IQ. Fundamentals of human neuropsychology. 7th editio. New York, NY: Worth Publishers; 2003.

4. Rowe FJ, Hepworth LR, Howard C, Hanna KL, Cheyne CP, Currie J. High incidence and prevalence of visual problems after acute stroke: An epidemiology study with implications for service delivery. PLoS One [Internet]. 2019 [cited 2019 Jul 2];14(3):e0213035. Available from: http://www.ncbi.nlm.nih.gov/pubmed/30840662

5. Làdavas E. The Role of Visual Attention in Neglect: A Dissociation between Perceptual and Directional Motor Neglect. Neuropsychol Rehabil. 1994;4(2):155–9.

6. Mancini F, Bricolo E, Mattioli FC, Vallar G. Visuo-haptic interactions in unilateral spatial neglect: The cross modal Judd illusion. Front Psychol. 2011;2(NOV).

7. Chatterjee A, Thompson KA, Ricci R. Quantitative analysis of cancellation tasks in neglect. Cortex. 1999;35(2):253–62.

8. Rowe FJ, Brand D, Jackson CA, Price A, Walker L, Harrison S, et al. Visual impairment following stroke: Do stroke patients require vision assessment? Age Ageing. 2009;38(2):188–93.

9. Edmans J, Lincoln N. The frequency of perceptual deficits after stroke. Clin Rehabil [Internet]. 1987 Nov 1;1(4):273–81. Available from:http://cre.sagepub.com/cgi/doi/10.1177/026921558700100403

10. Kerkhoff G. Neurovisual rehabilitation: Recent developments and future directions. J Neurol Neurosurg Psychiatry. 2000;68(6):691–706.

11. Jehkonen M, Ahonen JP, Dastidar P, Koivisto AM, Laippala P, Vilkki J, et al. Visual neglect as a predictor of functional outcome one year after stroke. Acta Neurol Scand [Internet]. 2000 Mar [cited 2016 Dec 8];101(3):195–201. Available from:http://www.ncbi.nlm.nih.gov/pubmed/10705943

12. Plante M, Demers L, Swaine B, Desrosiers J. Association Between Daily Activities Following Stroke Rehabilitation and Social Role Functioning Upon Return to the Community. Top Stroke Rehabil. 2010;17(1):47–57.

13. Mercier L, Audet T, Hébert R, Rochette a, Dubois MF. Impact of motor, cognitive, and perceptual disorders on ability to perform activities of daily living after stroke. Stroke. 2001;32(11):2602–8.

14. Kalra L, Smith DH, Crome P. Stroke in patients aged over 75 years: outcome and predictors. Postgrad Med J [Internet]. 1993 Jan 1;69(807):33–6. Available from: http://pmj.bmj.com/cgi/doi/10.1136/pgmj.69.807.33

15. Bazeley P. Qualitative data analysis with NVivo. SAGE; 2007. 217 p.

16. Krueger RA, Casey MA (Mary AW. Focus groups□;: a practical guide for applied research. SAGE; 2009. 219 p.

17. Strategyzer | Value Proposition Canvas [Internet]. [cited 2019 Jul 26]. Available from: https://www.strategyzer.com/canvas/value-proposition-canvas

18. Braun V, Clarke V. Using thematic analysis in psychology. Qual Res Psychol [Internet]. 2006 Jan;3(2):77–101. Available from: http://www.tandfonline.com/doi/abs/10.1191/1478088706qp063oa

19. Maxwell JA. Using Numbers in Qualitative Research. Qual Inq [Internet]. 2010 Jul 15;16(6):475–82. Available from:http://journals.sagepub.com/doi/10.1177/1077800410364740

20. Hepworth L, Rowe F. Ten Years On – A Survey of Orthoptic Stroke Services in the UK and Ireland. Br Ir Orthopt J [Internet]. 2019 May 8;15(1):89–95. Available from:http://www.bioj-online.com/articles/10.22599/bioj.135/

21. Hanna KL, Hepworth LR, Rowe FJ. Screening methods for post-stroke visual impairment: a systematic review. Disabil Rehabil [Internet]. 2016 Sep 26;1–13. Available from: https://www.tandfonline.com/doi/full/10.1080/09638288.2016.1231846

22. Kitsos G, Harris D, Pollack M, Hubbard IJ. Assessments in Australian stroke rehabilitation units: a systematic review of the post-stroke validity of the most frequently used. Disabil Rehabil [Internet]. 2011 Jan 9 [cited 2019 Aug 5];33(25– 26):2620–32. Available from: http://www.ncbi.nlm.nih.gov/pubmed/21554012

23. McCluskey A, Vratsistas-Curto A, Schurr K. Barriers and enablers to implementing multiple stroke guideline recommendations: a qualitative study. BMC Health Serv Res [Internet]. 2013 Dec 19 [cited 2019 Aug 5];13(1):323. Available from:http://www.ncbi.nlm.nih.gov/pubmed/23958136

24. Bouwmeester L, Heutink J, Lucas C. The effect of visual training for patients with visual field defects due to brain damage: A systematic review. J Neurol Neurosurg Psychiatry. 2007;78(6):555–64.

25. Hepworth LR, Rowe FJ. Visual Impairment Following Stroke - The Impact on Quality of Life: A Systematic Review. Ophthalmol Res An Int J [Internet]. 2016;5(2):1–15. Available from: http://sciencedomain.org/abstract/12864

26. Pollock A, Hazelton C, Brady M. Visual Problems After Stroke: A Survey of Current Practice by Occupational Therapists Working in UK Stroke Inpatient Settings. Top Stroke Rehabil. 2011;18(sup1):643–51.

27. Rowe FJ. Stroke survivors’ views and experiences on impact of visual impairment. Brain Behav [Internet]. 2017 Sep;7(9):e00778. Available from:http://doi.wiley.com/10.1002/brb3.778

28. NATIONAL INSTITUTE FOR HEALTH AND CARE EXCELLENCE (NICE). Stroke Rehabilitation: Long term rehabilitation after stroke [Internet]. NICE. Londo; 2013. Available from: http://www.nice.org.uk/nicemedia/live/14182/64094/64094.pdf

